# Maternal and perinatal health research during emerging and ongoing epidemic threats: a landscape analysis and expert consultation

**DOI:** 10.1101/2023.11.24.23298980

**Authors:** Mercedes Bonet, Magdalena Babinska, Pierre Buekens, Shivaprasad S Goudar, Beate Kampmann, Marian Knight, Dana Meaney-Delman, Smaragda Lamprianou, Flor Munoz-Rivas, Andy Stergachis, Cristiana M. Toscano, Joycelyn Bhatia, Sarah Chamberlain, Usman Chaudhry, Jacqueline Mills, Emily Serazin, Hannah Short, Asher Steene, Michael Wahlen, Olufemi T Oladapo

**Affiliations:** UNDP/UNFPA/UNICEF/WHO/World Bank Special Programme of Research, Development and Research Training in Human Reproduction (HRP), Department of Sexual and Reproductive Health and Research, World Health Organization, Geneva, Switzerland; Pharmacovigilance Team, Regulation and Prequalification Department (RPQ), Access to Medicines and Health Products Division (MHP), World Health Organization, Geneva, Switzerland; Department of Epidemiology, Tulane School of Public Health and Tropical Medicine, New Orleans, Louisiana, USA; KLE Academy of Higher Education and Research’s, J N Medical College, Belagavi, Karnataka, India; Charité Centre for Global Health, Universitätsmedizin Charité Berlin, Berlin, Germany; National Perinatal Epidemiology Unit, Nuffield Department of Population Health, University of Oxford, Oxford, UK; Division of Birth Defects and Infant Disorders, National Center on Birth Defects and Developmental Disabilities, CDC, Atlanta, Georgia, USA; Departments of Pediatrics and Molecular Virology & Microbiology, Baylor College of Medicine, and Texas Children’s Hospital, Houston, Texas, USA; School of Pharmacy and School of Public Health, University of Washington, Seattle, Washington, USA; Federal University of Goias, Institute of Tropical Pathology and Public Health, Goiania, Brazil; Boston Consulting Group, London, UK

## Abstract

**Introduction:** Pregnant women and their offspring are often at increased direct and indirect risks of adverse outcomes during epidemics and pandemics. A coordinated research response is paramount to ensure that this group is offered at least the same level of disease prevention, diagnosis, treatment, and care as the general population. We conducted a landscape analysis and held expert consultations to identify research efforts relevant to pregnant women affected by disease outbreaks, highlight gaps and challenges, and propose solutions to addressing them in a coordinated manner.

**Methods:** Literature searches were conducted from 1 January 2015 to 22 March 2022 using Web of Science, Google Scholar, and PubMed augmented by key informant interviews. Findings were reviewed and Quid analysis was performed to identify clusters and connectors across research networks followed by two expert consultations.

**Results:** Ninety-four relevant research efforts were identified. Although well-suited to generating epidemiological data, the entire infrastructure to support a robust research response remains insufficient, particularly for use of medical products in pregnancy. Limitations in global governance, coordination, funding, and data-gathering systems have slowed down research responses.

**Conclusion:** Leveraging current research efforts while engaging multinational and regional networks may be the most effective way to scale up maternal and perinatal research preparedness and response. The findings of this landscape analysis and proposed operational framework will pave the way for developing a roadmap to guide coordination efforts, facilitate collaboration, and ultimately promote rapid access to countermeasures and clinical care for pregnant women and their offspring in the future.

**Funding:** UNDP–UNFPA–UNICEF–WHO–World Bank Special Programme of Research, Development and Research Training in Human Reproduction, WHO, and Bill and Melinda Gates Foundation.

**Research in context:** *Evidence before this study:* Previous epidemics and pandemics highlighted the dearth of preparedness and response for maternal and perinatal health, resulting in access to countermeasures being delayed for this group, despite pregnant women and their offspring often being identified as at increased risk of severe disease outcomes. Based on this experience, we first searched PubMed from 1 January 2015 to 22 March 2022 with no language restrictions to identify any landscape analyses evaluating research efforts pertaining to pregnant women facing ongoing and emerging epidemic threats. Those efforts were defined as persistent data generation or aggregation exercises, including single studies, networks, and collaborations. As many of them struggled to secure and sustain baseline funding, it could be potentially beneficial to have them covered by some form of a global coordination mechanism to help improve their coherence. Multiple commentary articles discussing the need for harmonization of research and preparedness planning to avoid maternal and perinatal exclusion from potential preventative and treatment interventions in future epidemics/pandemics were identified, with most focusing on the lessons that can be learned from the COVID-19 pandemic. Evaluation of existing literature and scoping reviews identified studies which have evaluated gaps in approaches for alleviating gender inequality in future public health emergencies and the impacts of the COVID-19 pandemic on maternal and perinatal health services. None of them, however, have specifically focused on current research efforts in maternal and perinatal health that can be utilised in context of emerging and ongoing epidemic threats, or have proposed a framework for harmonizing future research efforts.

*Added value of this study:* This study provides a comprehensive overview of existing research efforts relevant to maternal and perinatal health in future outbreak, epidemic or pandemic situations. We summarise the key areas of focus of research efforts, identifying current gaps and areas in which the existing infrastructure is insufficient, and proposing an operational framework for improving conduct of maternal and perinatal heath research related to emerging and ongoing epidemic threats.

*Implications of all the available evidence:* The available evidence indicates that while current research efforts are well-suited to collecting maternal and perinatal epidemiological data, some gaps remain. They include limitations in global governance, coordination, funding, and data-gathering systems. The proposed operational framework developed based on the findings of this study will allow for development of a roadmap for guiding efforts and coordinating research to maximise access to countermeasures and clinical care for pregnant women and their offspring in during emerging and ongoing epidemic threats future outbreak, epidemic, and pandemic situations.

## Introduction

The likelihood of infectious disease outbreaks, epidemics, and pandemics is increasing and is expected to triple over the coming decades,^1^ due to a number of contributing factors such as increased travel, urbanization, and climate change.^2^ Historically, the emergence of epidemic-prone diseases, including Ebola, Zika, and respiratory infections such as Severe Acute Respiratory Syndrome (SARS), Middle East Respiratory Syndrome (MERS), and influenza A/H5N1 and A/H1N1 has caused global panic and alarm. However, disease emergence has often been followed by underinvestment in capacity strengthening, integrated surveillance, and protection of populations during the recovery phase.^3^ In this context, the ability to quickly gather information on the natural course of disease progression, clinical characteristics and pathophysiology is necessary for the development of prevention and clinical care strategies and guidelines, as well as for the planning, design, and delivery of care.^3^

Preparedness is key to reducing the impact of future disease outbreaks, and there are a number of lessons to be learned from the experience of the COVID-19 pandemic, where pre-pandemic response planning was limited, and handling of the health emergency at the global level was a considerable challenge.^4^ In the aftermath of the pandemic, the international community has called for strengthening of health emergency preparedness, response, and resilience (HEPR) architecture^1^ to better understand the distribution of priority emerging infectious diseases, together with drivers of transmission, natural history, clinical characteristics, disease pathophysiology, which can guide preparedness planning and strengthen health systems to ensure that they can effectively anticipate, respond to and recover from the impacts of any health emergencies.^3^ Integral to this response is the WHO R&D Blueprint, which brings together key stakeholders to identify gaps and accelerate research for accurate diagnostic assays, novel therapeutics, and effective vaccines against priority patogens.^5–7^

It is now globally recognised that a comprehensive research response to emerging and ongoing epidemic threats can and should contribute to better understanding on these affect the health and access to health care for women and children, in addition to their social and economic burden.^3^ Often, special populations, such as pregnant women and their offspring, are at higher risk both directly from the disease and from indirect factors. For example, pregnant women may be more likely to experience severe disease compared with non-pregnant women, as was noted during the COVID-19 and the 2009 influenza pandemics,^8, 9^ or their offspring may be at increased risk for developmental abnormalities, such as the association between microcephaly and maternal Zika infection observed during the 2015 outbreak in Brazil.^10, 11^ In addition to direct disease effects, pregnant women and their offspring are likely to be impacted by indirect effects, such as decreased access to maternity services, and increased childcare demands on working mothers during lockdown situations.^12, 13^ In addition, pregnant women are generally excluded from clinical trials of medicines and vaccines, resulting in delayed access to potentially life-saving treatments or preventative interventions.^14–17^

Our objective was to evaluate the current research landscape and identify major gaps and challenges to delivering a coordinated and rapid response for generating relevant scientific evidence to be better prepared for the next threat. We present the integrated findings of the landscape analysis, discussions with key informants (KIs) and outcomes of two expert consultations. We also propose an operational framework for maternal and perinatal research to be applied during ongoing and emerging epidemic threats.

## Methods

This landscape and gap analysis involved compilation and description of current research efforts relevant to maternal and perinatal health in the context of epidemics, and pandemics. As such, it formed the basis for a series of consultations to further identify main challenges and opportunities for coordination and generate ideas of how current research efforts could be leveraged to address gaps and develop an operational framework for improved maternal and perinatal health research during outbreaks. A Steering Committee was established to oversee and provide technical guidance at various stages of the project.

### Landscape and gap analysis

#### Desk review – search strategies and selection criteria

Initial searches were performed on Web of Science from 1 January 2015 to 22 March 2022 using three search strings including population (e.g., maternal/pregnancy), topic area (e.g., COVID-19, other infections), and methodology (e.g., various study designs) (See Supplementary Material 1). Research work undertaken by the authors identified in relevant literature was then further examined using Google Scholar and PubMed to find associated affiliations and additional publications. In parallel, a similar search was performed across grey literature, including governmental websites, relevant non-governmental and international organisations, conference proceedings, clinical trial registers, existing research effort websites and associated networks’ sites, as well as targeted Google searches.

A research “effort” was defined as a persistent data generation or aggregation exercise, which could be an individual study or a network or collaboration. Search results were filtered to exclude efforts considered to be beyond the scope of the study (e.g., only testing interventions in neonates) or focused on multi-year / lifelong longitudinal cohort studies’ or those that had otherwise been terminated. Broader efforts, such as the WHO Programme for International Drug Monitoring^18^ and ISARIC network^19^, were also excluded.

Preliminary findings from the literature review, grey literature and interviews were filtered against these screening criteria through manual review. The retrieved articles were screened by title and abstract to single out relevant full text documents to then evaluate them against the inclusion criteria. A data extraction form was used to extract information on the characteristics of those efforts (See Supplementary material 2), as well as opportunities and challenges pertaining to maternal and perinatal health research during epidemics and pandemics. The results were also cross-checked and tested via Quid analysis to address biases and cover blind spots. What remained at the end of the filtering process was included in the landscape analysis.

#### Preliminary contacts with key informants (KIs)

KIs were selected among the members of WHO Steering Committee, principal investigators or network members of efforts identified through the literature search. In total, twenty-three experts were contacted to identify further research efforts, gather more information on efforts led by KIs, and gain insights on opportunities and challenges for collaboration.

#### Quid analysis

Findings of the literature search described above were validated using Quid (Quid Inc., Business Intelligence Software, http://quid.com), an artificial intelligence software. This analysis allowed for finding gaps in the research landscape and clustering authors and research focuses and topics (e.g., Zika, birth & morbidity etc.) to detect networks and key individuals linking efforts, and to identify disparate, poorly linked clusters for which further investigation and outreach might be needed.

#### Synthesis of findings

Associated study publications, protocols, and websites were reviewed to determine the region where the effort was active, operational period (research duration), the population scope (e.g. maternal, neonatal, both or general population), type of research focus (e.g. observational, interventional, surveillance) and emergency focus (e.g., routine care or outbreak/epidemic). Results of desk research and expert interviews were used to evaluate research efforts and better understand the full scope of activities and related publications. When there was evidence of previous pandemic and epidemic-related work, emergency focus was included as part of the research scope. Furthermore, key networks of clusters and authors serving as connections were visually identified using Quid analysis. To gain additional understanding, a deeper characterization of selected efforts (exemplars) across a range of geographies and types was conducted. Key themes emerging from interactions with KIs were also identified and used to inform subsequent technical consultations.

Having explored how existing research platforms and research efforts could be leveraged to address gaps and how they could be used to meet the need for a comprehensive global emergency response, It was determined that structures and mechanisms would need to be established to approach dealing with new epidemics or pandemics in a holistic and coherent manner.

### Technical consultations

Two expert consultations were conducted in June 2022 and May 2023 to reflect on the results of the landscape analysis, learn from challenges and opportunities of exemplars, discuss an operational framework, and identify needs and next steps to produce concrete and actionable outputs for improved maternal and perinatal health research during epidemics and pandemics. A total of 33 attendees with broad expertise and relevant clinical and academic experience attended the meetings. Among them, 22 were women and 11 were men; 11 experts came from low- and middle-income countries (LMICs) while the remaining 22 experts represented high-income countries (HICs), most with direct experience in conducting or supporting research in Africa, Asia, and Latin America. In terms of the geographical representation of the WHO regions, three people came from Africa, 15 from the Americas, two from Eastern Mediterranean, ten from Europe, two from South-East Asia and one person came from the Western Pacific.

## Results

Overall, literature searches identified 3023 unique articles which were reviewed to identify relevant efforts corresponding with agreed definitions. Some articles yielded multiple efforts, while others yielded none. At the end of this process, a total of 94 research efforts remained all of which could be considered relevant for maternal and perinatal health research during future outbreaks (See Supplementary material 3). Studies that were beyond the scope of our research (e.g., only examining consumer product exposure among pregnant women in US) and those terminated prior to 2015 were excluded. The landscape analysis and expert consultations yielded three key findings leading to the development of an operational framework.

### Finding 1: Substantial research efforts exist, and there is likely already sufficient infrastructure to support robust maternal and perinatal health research during outbreaks

Multiple relevant research efforts are already in place. In total, 83% (78/94) of research efforts focused predominantly on both maternal and neonatal health (Figure 1a), with few efforts in the general population also including pregnant populations (2%, 2/94). These efforts have a broad geographical distribution, with 33% (31/94) being global efforts, 38% (36/94) originating from Europe or North America, and 29% (27/94) originating from the rest of the world (Figure 1b). Considerably fewer efforts were identified in Latin America, and there were no efforts solely based in the Eastern Mediterranean region. Data on duration were available for 81 research efforts, with the majority (60%) being operational for more than 5 years and 19% for more than 25 years (Figure 1c). Many of these efforts had been successfully utilised during the COVID-19 pandemic by leveraging existing protocols and clinical trials to collect data on COVID-19 burden, pregnancy outcomes, and use of medicines in pregnancy.^20–22^

**Figure 1.**
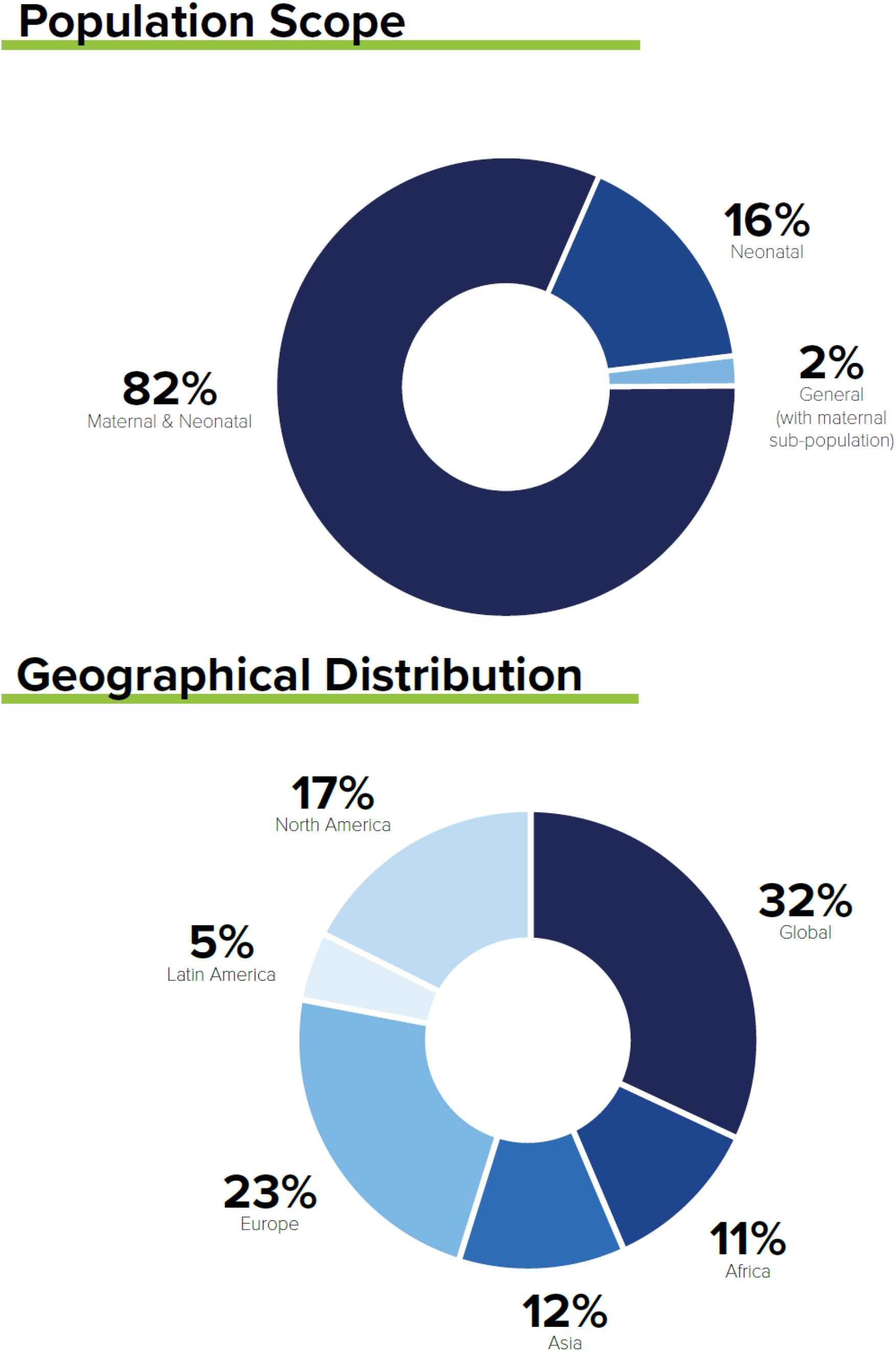
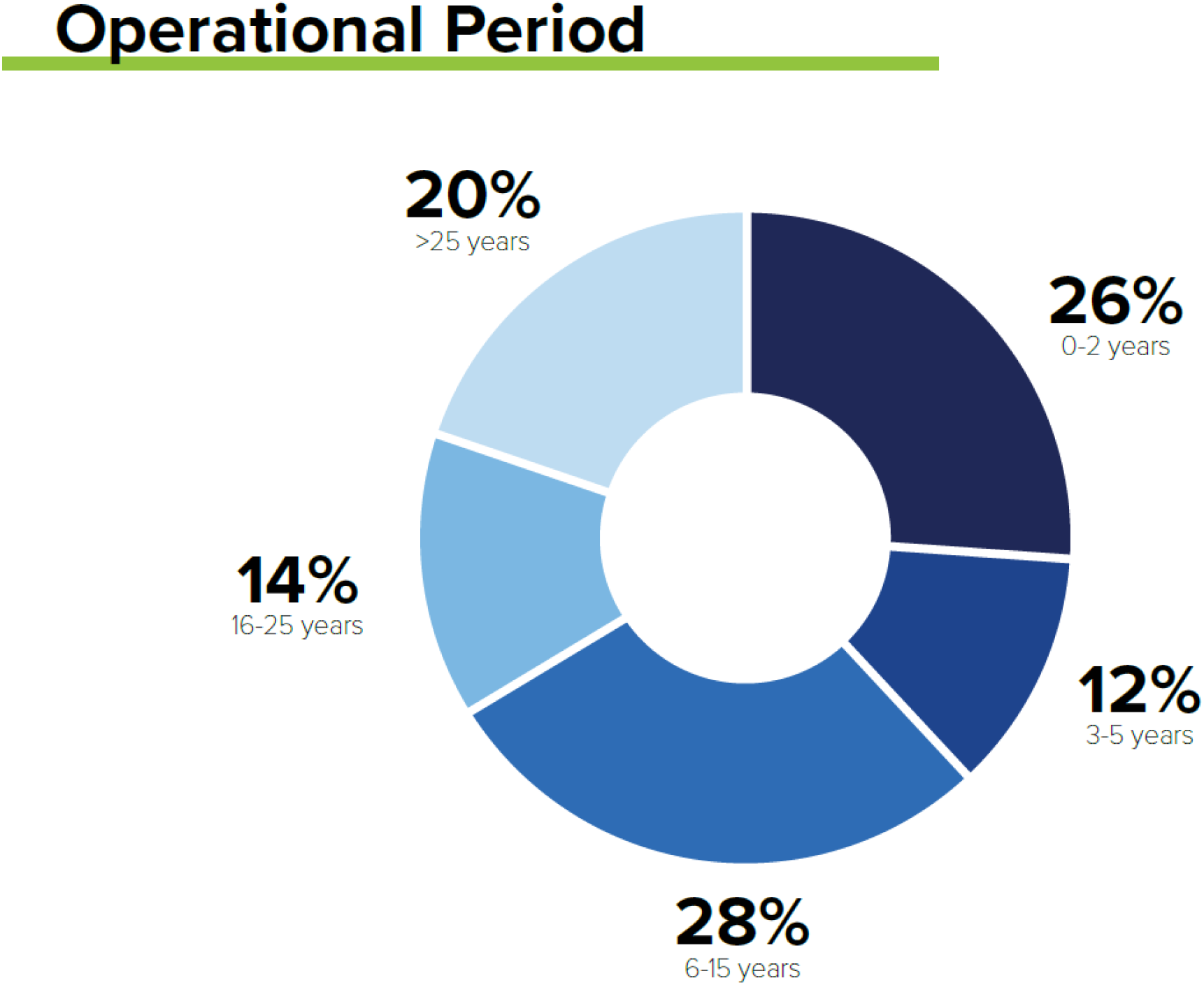
Characteristics of research efforts identified through literature searches and expert consultations.

Quid analysis showed that the over 3000 articles identified were authored by more than 20 000 researchers, in research networks consisting of more than 65 000 specific collaborations. Overall, the research ecosystem was predominantly comprised of discrete small clusters of research, with few connections (Supplementary Figure 1a). In total, 31 clusters included 0–5 authors, 10 included 6–10 authors, and 11 included >10 authors. The most connected network contained several of the largest clusters (Supplementary Figure 1b). The research landscape covered 16 major maternal and perinatal health topics, the most common being low-resource challenges (11%), diet and nutrition (11%), and vaccinations (12%; Supplementary Figure 1c).

Existing multi-country or regional networks may be the fastest path to improving maternal and perinatal health research during outbreaks. Large multi-country or regional networks already exist across epidemiology, research and development, post-marketing surveillance and advocacy. These existing networks may provide the quickest path to improving emergency preparedness. The International Network of Obstetric Survey Systems (INOSS; https://www.npeu.ox.ac.uk/inoss), the Global Network for Women’s and Children’s Health Research (https://globalnetwork.azurewebsites.net/),^23^ HIV/AIDS Clinical Trials Units and Clinical Research Sites (https://www.niaid.nih.gov/research/hivaids-clinical-trials-units-and-clinical-research-sites), and NEOCOSUR (https://neocosur.uc.cl/neocosur/vista/index.php) are already coordinating research and enabling collaboration on randomised controlled trials and cohort studies. Multi-site networks increase access to larger and more diverse study populations, which in turn increases the generalisability of study findings. In addition, alignment and coordination within networks can allow prompt cascading of new studies, protocols, or interventions to smaller satellite sites, which would not have been possible without cooperation within and among networks.

### Finding 2: Existing infrastructure is best suited to provide epidemiological data; research and development including pregnant women during outbreaks is limited

Approximately 87% of the identified efforts are suited to support rapid generation of epidemiological data, whereas only 9% focus on research and development of interventions. Many efforts conducted activities that contributed toward multiple categories (e.g., epidemiology and product development research).

Observational epidemiological efforts are suitable for rapidly leveraging the current infrastructure to describe the disease characteristics in outbreaks, including epidemics and pandemics. Existing efforts such as the UK Obstetric Surveillance System (UKOSS)^20^ and CONSIGN,^24^ the Global Network Maternal Newborn Health Registry,^23^ INTERCOVID^25^ and Ma-Cov^26^ were successfully adapted during the COVID-19 pandemic. Still, certain barriers remain, such as the speed of ability to amend existing protocols. Relatively few efforts focused on development of interventions, and the majority centred on repurposing existing efforts rather than introducing novel inventions. For example, excluding women from clinical trials resulted in a significant research gap during the COVID-19 pandemic,^14, 15^ although there were efforts those that advocated for improving inclusion of pregnant women in clinical trials (e.g., ConcePTION^27^). Furthermore, significant barriers to inclusion of pregnant women in clinical trials persist for developers of medicinal products, ranging from perceived higher levels of legal liability and reputational damage to unknown risks to the pregnant woman and the foetus. At the same time, relatively few incentives are available, despite the existence of guidance supporting inclusion of pregnant women in clinical trials.^16, 28, 29^

### Finding 3: Limitations in global governance, coordination and funding, and established data-gathering systems, cause delays in prompt, broad activation of response efforts during outbreaks

Establishing governance, coordination, and funding plans at the time, rather than in advance, of emergencies such as the Zika virus disease outbreak and COVID-19 pandemic delayed generation of evidence critical to determining the burden of disease and guiding public health and clinical management. For example, most of the maternal and newborn health efforts during the Zika outbreak occurred after cases had peaked, therefore missing critical periods for data collection and evidence generation for clinical decision-making. Efforts that required *de novo* development of studies and data gathering systems, including protocols, ethics approvals, data sharing agreements, etc., responded more slowly than those that had these structures in place. Studies which leveraged existing protocols and systems (e.g. UKOSS,^20^ Zika in Pregnancy in Honduras [ZIPH]^30^ and INTERCOVID^25^ studies) during the COVID-19 pandemic resulting in more rapid generation of epidemiological data compared to those studies developed and launched after COVID-19 had already emerged. These existing research efforts would benefit from increased global coordination, including harmonization of research protocols and pre-agreed data analysis plans and data-sharing agreements, to generate robust data that is applicable on an international scale. Specific funding to improve preparedness for pregnant women is not readily available, and many research efforts still struggle to obtain baseline funding. Funding for generating data concerning pregnant women is scarce and many research efforts are unable to secure and sustain baseline funding. It should be noted that prior to the COVID-19 pandemic, minimal investment was made for emergency preparedness and coordinated response, yet individual efforts (e.g., vsafe,^31^ UKOSS^20^) received funding for emergency response.

### Operational framework for maternal and perinatal health research during outbreaks

The operational framework (Figure 3) features three use cases (epidemiology, product development and post-authorization surveillance) that address the key gaps identified in the landscape review. The ability to generate epidemiological data on distribution, risks and burden of disease, and to facilitate its use for informed response and clinical guidance to enable prompt development of interventions is key. Conduct of trials that involve pregnant women where appropriate should support equitable development, access to and utilization of interventions. Properly conducted post-authorization surveillance activities would allow generation and communication of findings about the benefits and adverse effects of the use of medicinal products to further inform and update policy and practice.

**Figure 2.**
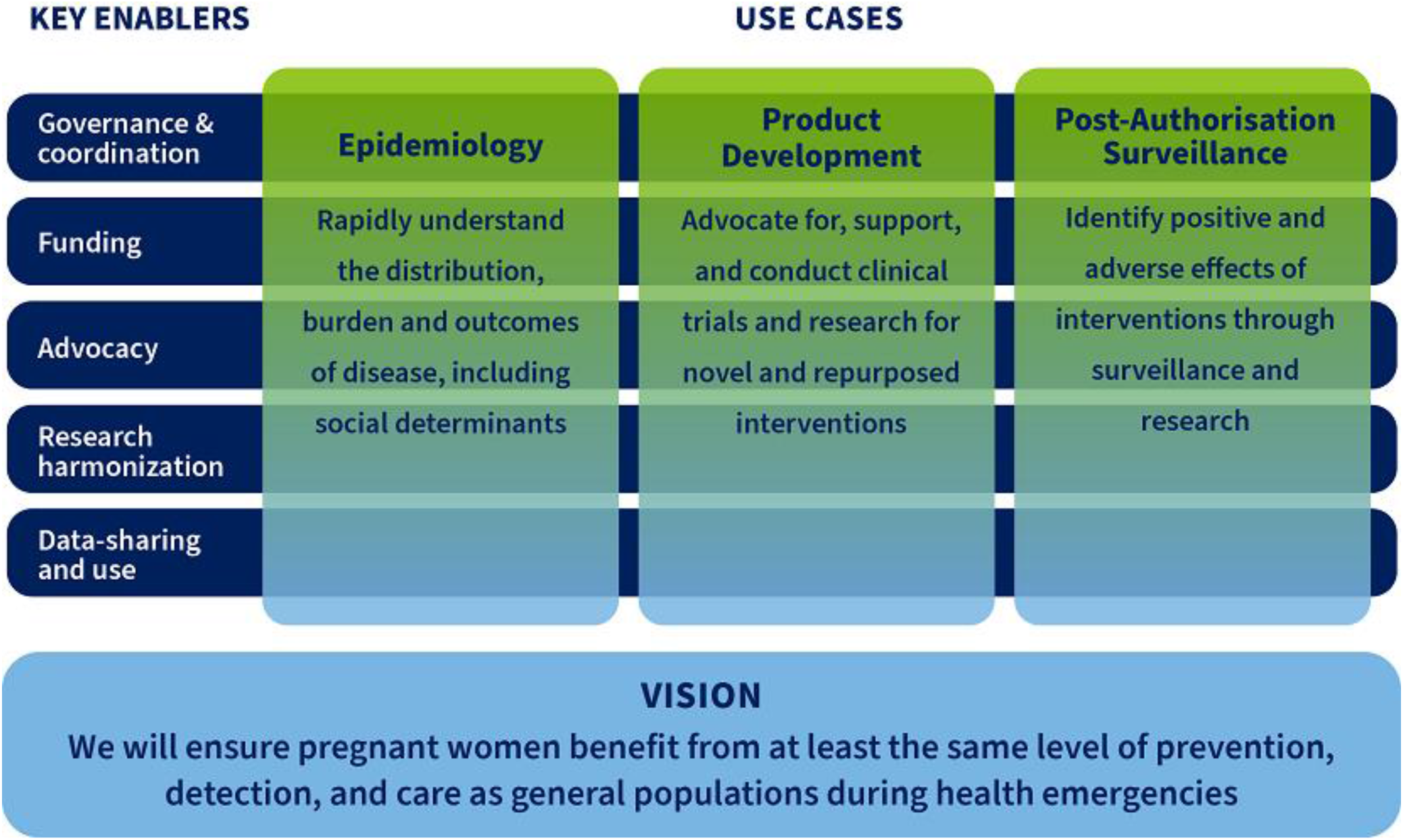
Operational framework for maternal and perinatal health research during outbreaks.

The three use cases supported by five key enablers (Governance and coordination, Funding, Advocacy, Research harmonization, Data-sharing and use) reflect interrelated actions that are needed to improve research efforts and decision-making related to pregnancy during ongoing and existing epidemic threats. The key enablers have been built on the needs identified during the two abovementioned expert consultations. Good governance and enhanced coordination mechanism are necessary to enable, direct, and oversee rapid research response encompassing research analyses and prompt dissemination of findings. Addressing health emergencies in a timely manner requires the presence of well-functioning sites, and a pool of trained personnel. Researchers in maternal and perinatal health should work hand in hand with public health administrators, policy makers and regulators on methods and data to be collected and shared in a manner that allows for informing policy and practice. The coordination mechanism should leverage existing platforms, ensuring that work is complimentary to and aligned with other preparedness initiatives directed at the general population. Establishing ‘centres of excellence’ or ‘sentinel sites’ should be supported as it would help close some of the existing gaps. Finally, opportunities ought to be created for research collaboration to continue at times when there are no outbreaks to maintain the existing infrastructure and promote continuous capacity building, particularly in low-resource settings.

In the initial phase of the project some funding would be required to establish major components of coordination and catalytic preparedness activities centred around capacity building, advocacy, harmonization, and data sharing and use. Incremental funding would help maintain research readiness and research implementation during outbreaks, encompassing data collection, publication and dissemination, and translation of findings into policy and recommendations. There is a need to map funding opportunities and proactively engage with donors to promote pre-agreed funding priorities and mechanisms.

Building and maintaining relationships with key stakeholders to encourage continuous interest in involving pregnant populations in research would be important enablers. Key stakeholders include researchers, health security and epidemiological surveillance actors, governments and policy makers, industry, regulators, patient groups, and civil society representatives, among others. High-profile advocacy is needed to remove barriers to research concerning pregnant women. Collaboration with pharmaceutical companies, who are often disincentivised from involving pregnant women in clinical trials, is needed to better understand and address their concerns. Underlining the ethical aspect of excluding pregnant women from research is essential as it could substantially help in facilitating the inclusion of pregnant women in trials, while encouraging the use of medicinal products in pregnancy, and disaggregation of epidemiological and surveillance data by pregnancy status. Another suggestion was to develop best practice guidance for community engagement and research, which would lead to effective engagement of women and civil society in epidemic and pandemic research. This covers efforts related to the dissemination of results and promotion of a proper uptake of medicines and vaccines once those have been proven to be effective. Advocating for the “general” pandemic funding to include sexual and reproductive health funding is advisable as it would serve to ensure that other emergency preparedness efforts launched in the wake of COVID-19 pandemic consider pregnant women’s needs.

Finally, equitable approaches should be used for development and implementation of research and data sharing, and to obtain relevant ethics and regulatory approvals. Harmonization of approaches, as opposed to complete standardization across sites, is highly desirable. It would enable rapid research response, minimizing delays to data collection and use, and influence policy and practice in a positive way. This entails development of harmonised research protocols for population-based epidemiological studies, clinical trials and post-marketing surveillance based on an agreed set of core variables/outcomes and definitions, including patient-centred outcomes, pre-agreed global data sharing principles, authorship rules and publishing principles. A mapping and analysis of existing protocols, data analysis plans and data sharing agreements would inform development of standard procedures applicable across different countries and networks in accordance with present consensus definitions such as the Global Alignment of Immunization safety, and other outcome sets and definitions for emerging diseases of epidemic and pandemic potential. This would serve to improve the availability of harmonised research tools and help streamline ethical review and approval processes while promoting data sharing and use by clinicians, regulatory authorities, policymakers and others.

## Discussion

This landscape analysis and consultative process identified 94 current research efforts applicable to maternal and perinatal research during emerging epidemic threats. It further supported developing a better understanding of limitations and challenges to deliver a more coordinated and rapid research response on pregnancy-related issues during outbreaks. Many gaps were identified, ranging from the need to cluster efforts towards epidemiological research to the need to scale up efforts related to research and development of medicinal products. In certain geographies, particularly in Latin America and Eastern Mediterranean, scarcity of research efforts was observed. Other regions suffered from lack of coordination, poor governance, insufficient funding, and limited harmonization of research and data sharing. An operational framework has been proposed to address all those gaps. It spans across three “use cases” (epidemiology, product development, and post-authorization surveillance) supported by five key enablers (governance and coordination, funding, advocacy, research harmonization, and data-sharing and use). The use cases would be ready for rapid deployment as per required geographic scope of an outbreak, thus allowing for a timelier decision making by policy makers, health workers and pregnant women themselves.

While some global efforts covered all regions, the analysis revealed clustering of research towards certain regions and specific use cases. Yet, despite recent outbreaks of Zika, chikungunya and dengue relatively few research efforts were found in Latin America and that is concerning. Similarly, few efforts were identified in the Eastern Mediterranean where MERS first appeared. Going forward, a well-designed research infrastructure should be established and maintained in all regions to generate data as soon as the need arises.

Ongoing efforts focused largely on collecting epidemiological data and relatively few efforts centred on product development or the logistics of product evaluation in pregnant women. Global collaborative research networks that use harmonised protocols and simplified data collection systems have accelerated the process of evidence generation.^32^ Maintaining and expanding these research networks will help accelerate the response to future emergencies.

Epidemiological efforts will be vital for providing data on risks and outcomes during ongoing and emerging epidemic threats, and informing development of clinical trials and post-marketing efforts that will encompass the population of pregnant women. Yet, additional engagement of stakeholders is desirable as it would help increase advocacy for inclusion of pregnant women in product development while allowing for a more rapid product delivery to this population in an emergency context. During the COVID-19 pandemic, a large number of clinical trials of selected vaccines and therapeutics systematically excluded pregnant women,^33^ while many of the products under evaluation had none or very low safety concerns during pregnancy.^14, 15^ Barriers for inclusion of pregnant women in trials persist, despite continuous calls for generation of efficacy and safety data during pregnancy in the context of outbreaks.^16, 33, 34^ The lack of clinical trial data in pregnant women hampers guideline development and public health advice. Frameworks and mechanisms defining when and how subpopulations such as pregnant women and children can and should be enrolled in clinical trials are needed to better address their needs. Currently, various guidance documents are being updated or developed at the international and national level, but none is specific to pregnancy research in the context of emerging and ongoing epidemic threats.

In contrast, effective networks and research studies are already under way or in place for conducting post-authorisation surveillance across many regions and they can be utilised during future outbreaks. Expanding them to cover additional geographies would provide a robust global picture of post-authorisation safety and allow for a rapid identification of any concerning signals in pregnant women or their offspring. The efforts potentially relevant to an emergency response identified in this landscape analysis fit into a broader landscape, which includes eight maternal and neonatal data collection systems in LMICs,^35^ over 170 pharmacovigilance organisations globally,^18^ and 52 clinical trial networks focused specifically on infectious diseases, including four in LMICs.^19^

Another gap that was identified referred to insufficient governance and lack of funding, leading to uncoordinated and slow responses during epidemics and pandemics. Established sites, trained personnel, and alignment among stakeholders are necessary for a coordinated emergency response which promotes inclusion of pregnant women in clinical trials, harmonises messaging, and achieves a maximal impact within the resources available. Outside of epidemic and pandemic situations, the framework provides the potential to expand research focusing on pregnant women and their offspring at the global and regional level, allowing for an increased focus on other maternal and child health priorities. In addition, it serves to promote greater collaboration among research groups and institutions resulting in co-publication of baseline data to be utilised by decision-makers as needed. Advocacy efforts underscore the importance of including pregnant women in clinical research, so that their needs are more likely to be considered in epidemic or pandemic situations. Stakeholder engagement is one of the key elements in achieving the vision of pregnant women benefitting from at least the same level of prevention, detection, and care as the general population during epidemics and pandemics. This maximises preparedness to ensure that this group would not be left behind in the future.

In summary, this landscape analysis and associated consultations identified numerous gaps that should be addressed to improve generation of data on maternal and perinatal health, and inform timely decision making by policy makers, health workers and pregnant women themselves, particularly in LMIC settings. Using an operational framework based on three use-cases and five supporting key enablers, the WHO/HRP aims to develop a roadmap to guide research into preparedness efforts, facilitate data consolidation to enable faster decision-making, and support readiness building, which covers harmonization of research protocols, data-sharing, and development of core outcomes to be collected.

## Supporting information

Landscape analysis

## Data Availability

All data produced in the present study are available upon reasonable request to the authors

## Abbreviations

HIC: high income country
HRP: Human Reproduction Programme
LMIC: low- and middle-income country
MERS: Middle East Respiratory Syndrome
SARS: Severe Acute Respiratory Syndrome
WHO: World Health Organization

## Contributors

MBo and OTO conceptualised the study; HS, JM conducted desk review; UC, MW conducted Quid analysis; MBo, MBa, OTO, SC, ES, AS, HS, JM coordinated the different expert consultations.

Members of the HRP Steering Committee (PB, SG, BK, MK, DMD, FMR, AS, CT) made significant contributions to the manuscript and facilitated preparations for the expert consultations in which they also participated. MBo and MBa drafted the manuscript with support from a medical writer. All authors provided comments and approved the final version of the manuscript.

## Data sharing

The data extracted and summarised in this scoping review are available upon request directed to the corresponding author.

## Acknowledgments

We gratefully acknowledge contributions of the participants to the in-person technical consultation of June 2022 and May 2023 (in an alphabetical order): Edgardo Abalos (Centro Rosarino de Estudios Perinatales), John Allotey (WHO Collaborating Centre for Global Women’s Health at the University of Birmingham), Mabel Berrueta (Institute for Clinical Effectiveness and Health Policy), Zulfiqar Bhutta (Hospital for Sick Children in Toronto, Canada), Robyn Churchill (USAID Bureau for Global Health), Amanda Cohn (Centers for Disease Control and Prevention), Stephanie Dellicour (Liverpool School of Tropical Medicine), Linda Eckert (University of Washington), Michelle Giles (Monash University), Zahra Hoodbhoy (Aga Khan University), Denise J. Jamieson (Emory University), Pisake Lumbiganon (Khon Kaen University), Charu Kaushic (Canadian Institutes of Health Research), Emeline Maisonneuve (Lausanne University Hospital), Anna Mastroianni (Johns Hopkins Berman Institute of Bioethics), Ushma Mehta (University of Cape Town), Edward Mullins (Imperial College London), Leslie Myatt (Oregon Health & Science University), Lisa Noguchi (Jhpiego), Saad Omer (Yale University), Aris T. Papageorghiou (University of Oxford), Helen Rees (University of Witwatersrand), Nadia A. Sam-Agudu (Institute of Human Virology Nigeria), Suzanne Serruya (Latin American Center of Perinatology, Women and Reproductive Health), Annick Sidibe (Jhpiego) Emily Smith (George Washington University), Stacie Stender (Jhpiego), Nathalie Strub Wourgaft (DNDi), Jimena Villar de Onis (Oxford University), and Gerald Voss (Coalition for Epidemic Preparedness Innovations-CEPI). Furthermore, we would like to acknowledge Bill and Melinda Gates Foundation (Ros Hollingsworth, Laura Lamberti, Anna Seale, Ajoke Sobanjo-ter Meulen); Boston Consulting Group (Joycelyn Bhatia, Sarah Chamberlain, Usman Chaudhry, Jacqueline Mills, Emily Serazin, Hannah Short, Asher Steene, Michael Wahlen); the HRP Scientific Committee: Pierre Buekens (Tulane University), Shivaprasad Goudar (Jawaharlal Nehru Medical College), Fyezah Jehan (Aga Khan University), Beate Kampmann (Charité Centre for Global Health), Marian Knight (University of Oxford), Dana Meaney-Delman (CDC), Flor Munoz-Rivas (Baylor College of Medicine), Andy Stergachis (University of Washington), Christiana Toscano (Federal University of Goias); WHO (Janet Diaz, Tracey Goodman, Sami Gottlieb, Edna Kara, Caron Rahn Kim, Smaragda Lamprianou, Vaseeharan Sathiyamoorthy, Shanthi Pal, Martina Penazzato, Diana Rojas Alvarez, Ronaldo Silva, Anna Thorson); WHO/HRP Secretariat (Magdalena Babinska, Mercedes Bonet, Olufemi Olapado). We would like to thank key informants who supported identification of further research efforts in the landscape analysis. Jenny Engelmoer (Sula Communications) is a medical writer and was contracted by Bill and Melinda Gates Foundation to draft the first version of the manuscript.

This study has received financial support from the UNDP–UNFPA–UNICEF–WHO–World Bank Special Programme of Research, Development and Research Training in Human Reproduction, Department of Sexual and Reproductive Health and Research, WHO, Geneva, Switzerland, and the Bill and Melinda Gates Foundation (INV-041181 WHO). The views of the funding bodies have not influenced the content of this manuscript.

## Supplementary materials

Supplementary Material 1: Search strategy

Supplementary Material 2: Data extraction form

Supplementary Material 3: Research efforts identified in desk review

Supplementary Figure 1: Maternal and perinatal health topics identified by Quid analysis

