## Supplementary material for "Maternal and perinatal health research during emerging and ongoing epidemic threats: a landscape analysis and expert consultation": Landscape analysis

### Supplementary Materials

**Supplementary Material 1. Search strategy in Web of Science**

TI = (maternal OR neonatal OR "neo-natal" OR "neo natal" OR pregnancy OR pregnant OR gestation* OR prenatal OR "pre natal" OR "pre-natal" OR perinatal OR "peri natal" OR "peri-natal" OR mothers OR "mother baby" OR "mother-baby" OR neonates OR lactat* OR "post-natal" OR "postnatal" OR "childbirth" OR "newborn")

AND

AB = (immuniz* OR vaccin* OR epidem* OR pandemic* OR endemic* OR "infectious" OR "nutrition" OR efficac* OR "influenza" OR "flu"OR coronavirus* OR covid OR "covid-19" OR zika OR ebola OR "SARS-CoV" OR "MERS-CoV")

AND

AB = (longitudinal OR "population-based" OR "population based" OR "population data" OR "cohort study" OR "cohort studies" OR surveillance* OR "post-marketing" OR "post marketing" OR "observational")

NOT

TI=(meta-analysis OR qualitative)

**Supplementary Material 2. Data extraction form**

| | |
|---|---|
| Site / network | [Name] |
| Links | [Name of PoC] |
| Brief Description | [Free text, covering background and aim of study] |
| Categorisation | [Type of study, emergency vs non-emergency focus, general vs maternal, interventional vs observational] |
| Scope | [Free text covering topics including disorders / diseases etc.] |
| Geography | [Countries and regions of surveillance] |
| Sample Size | [enrollment rate, # of pregnant women, deliveries registered etc.] |
| Lab capacity / capability | [Free text covering e.g., PCR capability, bio-safety levels etc.] |
| Protocols & mechanisms for amendments, including hard outcomes | [Free text covering e.g., detailed instructions on methodology, outcomes measured, reporting processes, cadence, identification and how to amend protocols etc.]<br>[Y/N against outcomes measured, including spontaneous abortion/miscarriage (10-19 wks gestation), stillbirth, preterm birth (<28wks, <32wks), low birth weight, NICU admission, maternal death, neonatal mortality (28 day)] |
| Other data priorities | [Free text] |
| Data collection | [Free text covering type of platforms, license & database, policies around data privacy & protection and definition of outcomes, + Y/N against ability to share and integrate data] |
| Funding & governance | [Free text covering sponsor / funding, institutions in charge of dev. / updates, cost for emergency research] |
| *Emergency response* | *[Free text covering any emergency response work currently conducted, and potential to pivot to emergency response in future]* |
| *Opportunities* | *[Free text covering any advantages of the site / network e.g. speed, practical use of data etc.]* |
| *Challenges* | *[Free text covering challenges e.g. funding for emergency response, comparison group, deidentified info etc.]* |

**Supplementary material 3. Research efforts identified in desk review**

| Effort (s) | Brief descriptions |
|---|---|
| AlignMNH | AlignMNH is a global initiative funded by the Bill & Melinda Gates Foundation in collaboration with the United States Agency for International Development. The initiative was founded in 2020 to accelerate progress in improving maternal and newborn health outcomes and prevent stillbirths around the globe. AlignMNH is supported by a Secretariat, managed by Jhpiego, an international NGO focused on transformative health care solutions that save lives. AlignMNH works in partnership with global and country-based organizations, national-level technical working groups, and other MNH-focused initiatives to more rapidly share science, evidence and programmatic experience across the maternal and newborn health communities |
| Alliance for Maternal and Newborn Health Improvement (AMANHI) Study | The Alliance for Maternal and Newborn Health Improvement (AMANHI) study in Sylhet district, Bangladesh, is using ongoing newborn intervention trials to obtain data critical to maternal, fetal and newborn health |
| Australasian Maternity OutcomesSurveillance System (AMOSS) | The Australasian Maternity Outcomes Surveillance System (AMOSS) is a national surveillance mechanism designed to study a variety of rare orserious conditions in pregnancy, childbirth and the postnatal phase in Australia and New Zealand. Through translating the findings from thesestudies into reliable evidence-based practice, the aim of AMOSS is to improve the safety and quality of maternity care in Australia and New Zealand |
| Australia and New ZealandNeonatal Network | The Australian and New Zealand Neonatal Network (ANZNN) is a collaborative network that monitors the care of high risk newborn infants by pooling data to provide quality assurance for this resource consuming care. The network was established in 1994 under the recommendation of the National Health and Research Council's (NHMRC) Expert Panel on Perinatal Morbidity. Since its establishment the network has developed a minimum data set and implemented a data collection that monitors the mortality and morbidity of infants admitted to neonatal intensive care units across Australia and New Zealand |
| Austrian obstetric surveillancesystem (AuOSS) | This is an obstetric surveillance system in Austria |
| Belgian Obstetrics Surveillance System (BOSS) | The Belgian Obstetric Surveillance System (B.OSS) was launched in 2011 with the support of the College of Physicians for Mother and Newborn of the Federal Public Service Public Health.B.OSS investigates serious complications that occur during pregnancy or childbirth and that put the life of the mother and/or the unborn child at risk. These are rare complications that affect less than one out of 2,000 women |
| BetterBirth Program | The BetterBirth Program is a project of the Ariadne Labs focused on ensuring better health and wellbeing for women, newborns, and infantsby improving quality and standards of care, minimizing complications, and ending preventable deaths. They do this through the implementation of scalable, evidence-based solutions that work in the real world, both at the frontline of care and in communities. The BetterBirth Program began with the goal of evaluating the World Health Organization’s (WHO) Safe Childbirth Checklist and identifying opportunities and strategies to support its successful implementation. Since that time, it has grown to address the complexities across the ecosystem of maternal, newborn, and infant health |
| Brazilian Obstetric Observatory (OOBr) | The objective of this project is to create an obstetric observatory through an interactive platform for monitoring, analyzing public data and disseminating information in the area of Obstetrics in Brazil. It will provide exploratory data analysis with the purpose of assessing the |

| | |
|---|---|
| | impacts of the H1N1 (2009) and COVID-19 (2020) pandemics on maternal, fetal and neonatal health |
| Canadian Neonatal Network | This is a group of researchers who collaborate on research issues relating to neonatal care with members from 30 hospitals and 17 universities. It maintains a standardized neonatal intensive care unit (NICU) database and provides a unique opportunity for researchers to participate in collaborative projects on a national and international scale; supports clinical, epidemiologic, outcomes, health services, health policy and informatics research aimed at improving efficacy and efficiency of neonatal care |
| CHAMPS | The Child Health and Mortality Prevention Surveillance Network (CHAMPS) was established to develop a network of high-quality sites to collect robust and standardized longitudinal data, with the overarching objective of understanding and tracking the preventable causes of childhood death globally. |
| CMC Vellore | They have a surveillance study that collects risk factors and outcomes of pregnant women every 4 months via cross sectional surveys, home visits and a longitudinal study that investigates data on key maternal and infant health indicators. |
| ConcePTION project | ConcePTION is a 5-year program by funded by the Innovative Medicines Initiative. It is a research consortium aimed at establishing an international network and framework to improve the evaluation of the safety of medicines in pregnancy and breastfeeding. It represent 88 public and private organisations |
| Coronavirus Health Outcomes inPregnancy and Newborns | The CHOPAN registry aims to collect real-time data on pregnant women who are infected with the coronavirus that causes COVID-19 (SARS-CoV2) to improve understanding of its impact on pregnancy outcomes. This registry provides regular feedback to clinicians and public health officials to allow evidence-based management of women and their babies with coronavirus infection. This registry was developed by Australian clinicians in collaboration with international colleagues in order to maximise its value to the global community |
| COVID-19 in pregnancy(PregCOV-19LSR) | The PregCOV-19 project aims to undertake living systematic reviews (LSR) involving pregnant and postnatal women at risk, suspected, and diagnosed to have COVID-19, and synthesise the relevant evidence on prevalence, risk factors, mother-to-child transmission, diagnosis, treatment of the disease. The findings will be continuously updated, by incorporating appropriate new evidence as it becomes available |
| COVID-19 Vaccines International Pregnancy Exposure Registry (C-VIPER) | The objective of the COVID-19 Vaccines International Pregnancy Exposure Registry (C-VIPER) is to evaluate obstetric, neonatal, and infant outcomes among women vaccinated during pregnancy with a COVID-19 vaccine. They measure obstetric outcomes (spontaneous abortion, antenatal bleeding, gestational diabetes, gestational hypertension, intrauterine growth restriction, postpartum hemorrhage, fetal distress, uterine rupture, placenta previa, chorioamnionitis, Caesarean delivery, COVID-19), neonatal outcomes (major congenital malformations, low birth weight, neonatal death, neonatal encephalopathy, neonatal infections, neonatal acute kidney injury, preterm birth, respiratory distressin the newborn, small for gestational age, stillbirth, COVID-19), and infant outcomes (developmental milestones [motor, cognitive, language, social-emotional, and mental health skills], height, weight, failure to thrive, medical conditions during the first 12 months of life, COVID-19) among pregnant women exposed to single (homologous) or mixed (heterologous) COVID-19 vaccine brand series from 30 days prior to the first day of the last menstrual period to end of pregnancy and their offspring relative to a matched reference group who received no COVID-19 vaccines during pregnancy |
| COVID-19-Associated Hospitalization Surveillance Network (COVID-NET) | The COVID-19-Associated Hospitalization Surveillance Network (COVID-NET) collects data on hospitalized pregnant women with laboratory-confirmed SARS-CoV-2, the virus that causes COVID-19; to date, such data have been limited |

| | |
|---|---|
| COVID-19-Related Obstetric and Neonatal Outcome Study (CRONOS) | COVID-19 Related Obstetric and Neonatal Outcome Study (CRONOS) is an ongoing prospective multicenter registry study for SARS-CoV-2 positive pregnant women, initiated on 3 April 2020 by the German Society of Perinatal Medicine |
| Delivery Decision Initiative | The Delivery Decisions Initiative is a project from the Ariadne labs, conducting research and social impact work, focused on transforming childbirth care around the world so that every person can start or grow their family with dignity. They develop and implement efforts aimed at improving equitable maternal health care via collaborations with health systems and communities, designing systems that are capable of producing safe, supportive and empowering care for every birthing person everywhere |
| EUROCAT | European system for aggregating information from registries gathering data on congenital anomalies and pregnancy outcomes (livebirths, stillbirths and terminations of pregnancy), updated twice a years |
| German Obstetric SurveillanceSystem (GerOSS) | GerOSS (German Obstetric Surveillance System) aims at generating deeper insight into relevant risk factors to improve diagnosis and treatment of severe complications during pregnancy and delivery. As such it is primarily conceived as a system for quality improvement and less as a register. Another focus is the provision of an information and communication platform for dissemination of these insights. Finally, incidences of selected rare obstetric events may be derived |
| Global Network Maternal Newborn Health Registry (GN-MNHR) (includingCOVID study) | This is an active maternal and neonatal data collection system with multiple LMIC sites. A prospective, population-based registry that startedin 2008; it enrolls and follows up on pregnant women and their newborns up to 42 days postpartum. A sub-study to the Maternal Newborn Health Registry will be conducted to understand the prevalence of COVID-19 among pregnant women, the association between COVID-19 and pregnancy outcomes, and the Knowledge, Attitudes, and Practices of pregnant women related to COVID-19 and its prevention during pregnancy. Women who consent to participation in the MNHR COVID sub-study will provide a blood specimen at or near delivery to be tested for COVID-19 antibodies. Four Global Network sites (Guatemala, Bangladesh, Nagpur India and Pakistan) will also collect and analyze blood specimen from participants during antenatal care visits |
| Global Network Maternal NewbornHealth | The Global Network for Women's and Children's Health Research (Global Network) is a partnership dedicated to improving maternal and child health outcomes and building health research capacity in resource-poor settings by testing cost-effective, sustainable interventions that provide guidance for the practice of evidence-based medicine. This unique collaboration includes 8 multidisciplinary research sites and a Data Coordinating Center. Scientific oversight for the Global Network comes from the Eunice Kennedy Shriver National Institute of Child Health and Human Development, which is part of the National Institutes of Health within the U.S. Department of Health and Human Services |
| Global Pregnancy Collaboration (CoLab) | The goal of the Global Pregnancy Collaboration (CoLab) is to improve the health of mothers and their infants by facilitating harmonized perinatal data management and collaborative research. This is accomplished by developing standardized generic data dictionaries that cover the needs of all maternal child health care—from the simplest in the most resource poor countries to the most sophisticated research in moreprivileged areas |
| Grupo Castrillo- Spanish NeonatalNetwork | Grupo Castrillo is a Spanish network for neonatal infections surveillance. Since 1995 the network is collecting prospective data about neonatal sepsis and antimicrobial resistance, viral infections, congenital infections, and other related topics (infectious biomarkers, central catheter complications, etc.) |
| Helping Mothers Survive and Helping Babies Survive | Jhpiego participates in a consortium of global partners to provide and promote these training programs that provide evidence-based learningmodules designed to improve and sustain the skills of midwives, nurses, doctors, and those who provide direct care during pregnancy, labor and birth. |

| | |
|---|---|
| Interagency working group onreproductive health in crises | IAWG is an international coalition of organizations and individuals working collectively to advance sexual and reproductive health and rights in humanitarian settings |
| INTERCOVID study | This is the next phase of the INTERGROWTH 21ST project. It is a study aimed to develop new "prescriptive" standards describing normal fetal growth, preterm growth and newborn nutritional status in eight geographically diverse populations, and to relate these standards to neonatalhealth risk (INTERBIO 21ST) |
| International Clearinghouse for BirthDefects Surveillance and Research | The organization brings together birth defect surveillance and research programmes from around the world with the aim of investigating andpreventing birth defects and lessening the impact of their consequences |
| International Network for Evaluation of Outcomes of Neonates | iNeo maintains a standardized neonatal intensive care unit (NICU) database and provides a unique opportunity for researchers to participate in collaborative projects on a national and international scale. Health care professionals, health services researchers and health administrators participate actively in clinical and epidemiological outcomes, health services, health policy and informatics research aimed at improving the efficacy and efficiency of neonatal care |
| International Network of ObstetricSurvey Systems (INOSS) | INOSS is a multinational collaboration of organizations conducting prospective population-based studies of serious illnesses in pregnancyand childbirth |
| ISGlobal Maternal, Child, Reproductive Health Research | The Barcelona Institute for Global Health (ISGlobal) was set up in 2010 as a result of an initiative of the "la Caixa" Foundation working with academic and government institutions interested in creating a centre of excellence in research and knowledge translation in Barcelona equipped to meet the new challenges facing global health in the 21st century. Our roots, however, go back much further. Today, ISGlobal encompasses over 30 years' experience in the field of health and is a consolidated hub of excellence in scientific research drawing on expertise from both the hospital environment and academic institutions |
| Israeli Neonatal Network | The Israel Neonatal Network (INN) is a voluntary consortium of all neonatal departments in Israel. The Israel National Very Low Birth Weight VLBW) Infant Database was established in 1995 under the auspices of the INN. The main objectives of the database are the application of quality data for the assessment of morbidity and mortality trends of VLBW infants; benchmarking of individual neonatal unit performance in comparison to national data; quality of care and management; planning of national, regional and institutional structure and policy development; longitudinal developmental assessment and for collaborative research programs |
| Italian Obstetric Surveillance System(ITOSS) | The Italian Obstetric Surveillance System (ItOSS) collects and disseminates information on severe maternal morbidity and mortality. Since 2017, the surveillance system of maternal mortality, coordinated by the Istituto Superiore di Sanità (ISS), has been collecting comprehensive and reliable data on maternal mortality in 13 Italian regions (Piedmont, Lombardy, Veneto, Friuli Venezia Giulia, Emilia-Romagna, Marche, Tuscany, Lazio, Campania, Apulia, Calabria, Sicily and Sardinia), accounting for 91% of total births in Italy. Between 2008 and 2016, through the National Centre for Disease Prevention and Control (Centro nazionale per la prevenzione e il controllo delle malattie – CCM), the Ministry of Health consistently funded a series of ISS-coordinated multiregional projects which allowed further development and consolidation of the surveillance |
| JHU International Center for Maternaland Newborn Health | This interdisciplinary team applies expertise in public health, behavioral sciences, nutrition, medicine, engineering, and informatics. They rely on workforce development in Sub-Saharan Africa and South Asia to implement and evaluate low-cost solutions to prevent illnesses and deaths in the world's most vulnerable mothers and babies |
| Korean Neonatal Network (KNN) | A neonatal surveillance network in Korea |

| | |
|---|---|
| Le Comité national d'experts sur la mortalité maternelle (CNEMM) | The National Committee of Experts on Maternal Mortality (CNEMM) was created in 1995 by order of the Ministry of Health with the mission ofexamining maternal deaths documented by a confidential investigation, identifying the factors involved in the occurrence of these deaths and to propose preventive measures. This mission involves a specific information collection system, the National Confidential Survey on Maternal Mortality (ENCMM), whose scientific coordination is ensured by the National Institute of Health and Medical Research (Inserm) (unit 1153 , EPOPé team) |
| Magee Obstetric Maternal and Infant(MOMI) Database and Biobank | MOMI Database and Biobank collects obstetric biological materials along with annotated clinical information via a rigorous process for qualitycontrol. Our unique, web-based inventory system tracks our biological specimens, linking them to annotated clinical data behind a secure, HIPAA compliant server |
| Malawi Maternal Health and Safe Motherhood Initiative | This is a maternity waiting village in malawi, with support from UNC Global Women's health. They have several ongoing studies, several focusing on HIV |
| Maternal health and Covid-19(MA-CoV) Study | Ma-CoV is a study aimed to characterize the clinical presentation of COVID-19 in pregnancy, evaluate the incidence of infection during pregnancy, identify risk factors of maternal and neonatal morbidity and mortality associated with SARS-CoV-2 infection as well as the risk of mother-to-child transmission of SARS-CoV-2 |
| Maternal health task force | The Maternal Health Task Force (MHTF) at the Harvard Chan School of Public Health strives to create a strong, well-informed and collaborative community of individuals focused on ending preventable maternal mortality and morbidity worldwide. The vision for MHTF is that it serves as a space that not only identifies and shares promising research, but serves as a catalyst for research improvement and innovation |
| Maternal, Newborn & Child HealthWorking Group (MNCH-WG) | THis is an active maternal and neonatal data collection system. INDEPTH is a network of independent Health and Demographic Surveillance System (HDSS) sites that carry out longitudinal research. The INDEPTH Network Maternal, Newborn & Child Health Working Group (MNCH-WG)coordinate the surveillance of pregnancies and outcome tracking |
| Maternal-fetal medicine units (MFMU) network at UPMC Magee-Womens Hospital | NICHD established the MFMU Network in 1986 to respond to the need for well-designed clinical trials in maternal-fetal medicine and obstetrics, particularly with respect to preterm birth. The aims of the network are to reduce maternal, fetal, and infant morbidity related to preterm birth, fetal growth abnormalities, and maternal complications and to provide the rationale for evidence-based, cost-effective obstetric practice |
| MATIMMUNE Study | The main objective of this study is to investigate the impact of timing of vaccination during pregnancy on humoral immune responses in pregnant women at several timepoints during and after pregnancy |
| Medical Birth Registry of Norway | All maternity units in Norway must notify births to the MBRN. The notification form includes the name and personal identity number of the child and parents, as well as information about maternal health before and during pregnancy, and any complications during pregnancy or birth. This includes information about medicine use in pregnancy, labour interventions, birth complications, maternal complications after birth, whether the baby is born alive, any diagnoses in the child or evidence of congenital abnormalities |
| MNCH Morbidity and MortalitySurveillance - HaSET | A prospective longitudinal cohort study of pregnant women and children less than two years of age with an aim of understanding the epidemiology of maternal and childhood illnesses and deaths in selected Kebeles of Angolela Tera, Kewet and Shewarobit Woredas in North Shewa Zone, Amhara Region, Ethiopia. |

| | |
|---|---|
| Momentum Maternal and Perinatal Death Surveillance and Review (MPDSR) | MOMENTUM Country and Global Leadership, the World Health Organization, UNFPA, and UNICEF have developed an integrated Maternal andPerinatal Death Surveillance and Response (MPDSR) Capacity Building Package that can be used to support country capacity for MPDSR through virtual means. Capacity building materials for both maternal and perinatal mortality surveillance have been combined into one package, which also includes updated guidance and COVID-19 modules |
| MotherToBaby | MotherToBaby Pregnancy Studies provide information on medication and vaccine safety in pregnancy. Studies are observational; people who are pregnant are not asked to take any medications or change their current treatments. They follow people who are pregnant who have – andwho have not – taken a medication of interest until they deliver their baby, and then follow their babies for a period of time after birth. They collect information along the way that allows determination of whether the medication/vaccine may pose a risk to a pregnancy or a developing baby |
| MPD-4-QED (Nigeria) | This program was initiated in Nigeria in 2019 across 54 tertiary level public and private referral level health facilities. It was established to implement a standardized electronic platform for the collection and collation of maternal and perinatal data, to enable routine healthcare data analysis and maternal and perinatal death audits. It will facilitate reporting and feedback at the local, state, regional, and national levels for the improvements in clinical care performance. Currently in its third year with >200,000 participants enrolled. |
| Multi-Omics for Mothers and Infants(MOMI) Biorepository Platform | This platform builds off an existing pregnancy biobank in the Sylhet district of Bangladesh, known as AMANHI-Bangladesh. The AMANHI-Bangladesh biobank contains biological samples from mothers and infants and related clinical and epidemiological data. Researchers use the sample and data collected by the biobank to identify biological and genetic markers that may predict a mother's increased risk of adverse outcomes including preeclampsia, preterm birth, and small for gestational age (smaller or less developed than normal for the baby's sex and gestational age). Through the MOMI Biorepository Platform grant, researchers will maintain the existing biobank infrastructure and develop a scale-up plan that will include other biobank sites and partners. Researchers also aim to identify new insights and biomarkers for preterm birth and other pregnancy complications that will help to inform predictors and treatment options. The MOMI Biorepository Platform will alsoinclude capacity-building components for research staff and data scientists at the Bangladesh field site |
| National Neonatal ResearchDatabase (NNRD) | The NNRD is a national resource holding real-world clinical data captured in the course of care on all admissions to NHS neonatal units in England, Wales, Scotland and the Isle of Man. Neonatal units submit data through their Electronic Patient Record system supplier. At present, there is information on around one million babies and 10 million days of care in the NNRD. The NNRD is available to support audit, evaluations, bench-marking, quality improvement and clinical, epidemiological, health services and policy research to improve patient care and outcomes |
| National Registry for surveillance and epidemiology of perinatal Covid-19 infections | This National Registry represents a collaboration between the American Academy of Pediatrics Section on Neonatal-Perinatal Medicine, the Vermont-Oxford Network (VON) and MedNAX (an organization of private neonatologist)h |
| NEOCOSUR network | This network includes 36 public and private tertiary centers, all University-affiliated. They aim to conduct continuous evaluation of mortality and morbidity of very low birth weight infants (VLBWI) population in the region, develop of predictive adjustment tools that allow benchmarking between centers, conduct clinical observational studies with data provided from the DBU, and non-interventional prospective studies, and design/ conduct clinical trials to evaluate the effectiveness of specific therapeutic interventions |

| | |
|---|---|
| Neonatal Research Network Japan | This is the Neonatal research network database in Japan. All infants who were born in the participating hospitals with gestational age less than 32 weeks and with birth weight at or less than 1500g admitted to participating facilities within 28 days after birth are registered in the database. Those infants who were born alive but died in a delivery room are also included. As of January 1, 2021, 131 perinatal centers are participating in the network |
| Nordic Obstetrics SurveillanceSystem (NOSS) | This is an obstetric surveillance system collecting data from the Nordic countries |
| Obstetric-fetal pharmacology researchcenter (OPRC) | The mission of the OPRC Network, formerly the Obstetric-Fetal Pharmaceutical Research Units Network, was to improve the safety and effective use of therapeutic drugs in pregnant and lactating people. The network's overall goal was to promote and facilitate cooperative multidisciplinary research to enhance the understanding of obstetric pharmacokinetics (PK) and pharmacodynamics (PD). OPRCs provided theexpert infrastructure needed to test therapeutic drugs during pregnancy. The centers allowed researchers to conduct safe, technically sophisticated, and complex studies that helped clinicians protect the health of pregnant people, improve birth outcomes, and reduce infant mortality |
| PAHO Perinatal InformationSystem (SIP) | This is an active maternal and neonatal data collection system. Perinatal Informatic System (SIP) by PAHO is a perinatal clinical record that Ministries of health and maternity services (public and private) have adopted |
| peri-COVID | The periCOVID study was set up by a group of doctors and researchers who are interested in understanding if pregnant women who test positive for the novel coronavirus (SARS-COV-2) can transmit the infection to their unborn babies and if baby's of mothers who have been vaccinated against SARS-CoV-2 will be protected from COVID-19 infection |
| Perinatal Problem IdentificationProgramme | The Perinatal Problem Identification Program (PPIP) is a tool that makes perinatal and maternal death audits easier. It provides simple analysis on monthly deaths, causes of death and avoidable factors |
| Pregnancy and Neonatal Outcomes inCOVID-19 (PAN-COVID) | PAN-COVID is a global registry of women with suspected COVID-19 or confirmed SARS-CoV-2 infection in pregnancy and their neonates; understanding natural history to guide treatment and prevention. The overall aims of PAN-COVID are to evaluate the association of suspectedCOVID-19 and confirmed SARS-CoV-2 infection in women in pregnancy with: 1. Miscarriage, 2. Fetal growth restriction and stillbirth, 3. Pre- term delivery, 4. Vertical transmission. They collaborate with 13 other national and international COVID-related maternal and neonatal registries |
| Pregnancy Risk Assessment Monitoring System (PRAMS) | PRAMS, the Pregnancy Risk Assessment Monitoring System, is a surveillance project of the Centers for Disease Control and Prevention (CDC) and health departments. Developed in 1987, PRAMS collects jurisdiction-specific, population-based data on maternal attitudes and experiences before, during, and shortly after pregnancy. PRAMS surveillance currently covers about 81% of all U.S. births |
| PRiSMA | PRiSMA is a network of sites funded by BMGF collecting anthropometric, sociodemographic, pregnancy, and child health data. They conduct epidemiological studies, but are also developing the capacity to conduct clinical trials in their populations |
| QoC WHO | The goal of the Quality of Care is to work with the WHO to develop national quality and operational plan to reduce maternal and neonatal mortality. QoC programmes have been set up in Bangladesh, Cote d'Ivoire, Ethiopia, Ghana, India, Kenya, Malawi, Sierrra Leone, Tanzania, Uganda |

| | |
|---|---|
| Registry for pregnant women exposedto SARS-CoV-2 (COVID-19, CONSIGN study) | The COVI-PREG registry aims to collect data to understand the natural history of the SARS-CoV-2 among pregnant women and the impact on maternal, pregnancy and neonatal outcomes. The study objectives are to characterize the clinical course of SARS-CoV-2 infection during pregnancy, assess the risk assessment of vertical transmission and congenital lesions, quantify the risk of adverse maternal outcomes, pregnancy outcomes and neonatal outcomes, and identify additional risk factors and risk modifiers |
| Reproductive, Maternal, & Child Health (RMNCH) | This is a collaboration of a Nepalese and US-based NGOs, supporting health innovation in Nepal, conducting research and innovation to address evidence, implementation, and policy gaps in the equity, quality and accessibility of healthcare. They have a particular MNCH research division |
| Reproductive, Maternal, Newborn, Child, and Adolescent Health (RMNCAH)division - Impact of COVID-19 on the Health and Nutrition of Women and Children in Low- and Middle- Income Countries study | A hub for global child health-focused activities and connects researchers and health-care professionals around the world, with focus on RMNCAH; Special interests in scaling up evidence-based, community setting interventions and implementation of reproductive, maternal, newborn, child and adolescent health and nutrition interventions in humanitarian contexts |
| Safety Platform for EmergencyVaccines (SPEAC) | This effort is funded by CEPI to promote harmonization of safety assessment across research platforms and enable meaningful analysis and interpretation of the safety profile of CEPI vaccines |
| Sanofi Pasteur PregnancySurveillance Program | The Sanofi Pasteur Pregnancy Registries are an organized, systematic collection of data on pregnant women vaccinated with one or more of the following vaccines: Menactra® (Meningococcal [Groups A, C, Y and W-135] Polysaccharide Diphtheria Toxoid Conjugate Vaccine), Adacel®(Tetanus Toxoid, Reduced Diphtheria Toxoid and Acellular Pertussis Vaccine Adsorbed), Fluzone® Quadrivalent (Influenza Virus Vaccine), MenQuadfi® (Meningococcal [groups A, C, Y, W] conjugate vaccine), Dengvaxia®(Dengue tetravalent vaccine, live, attenuated), Flublok® Quadrivalent (Influenza Virus Vaccine) |
| SASOG - Covid 19 study | The primary aim of the present study was to describe the char-acteristics and outcomes of hospitalized pregnant women infected with SARS-CoV- 2 in South Africa who were admitted for treatment of clinical SARS-CoV- 2 illness or other indication, in order to inform evidence-basedguidance for pregnant women in South Africa |
| SET-NET | CDC's Surveillance for Emerging Threats to Mothers and Babies Network (SET-NET) detects the effects of health threats on pregnant people and their babies by collecting data from pregnancy through childhood. It uses evidence-based, actionable information to help save and improve the lives of mothers and babies |
| Shoklo Malaria Research Unit - Motherand child health team | The mother and child health (MCH) team run a network of antenatal clinics and delivery facilities for the border population and have documented over 70,000 pregnancies and their outcomes. This massive effort has resulted in a phenomenal contribution to the evidence base on the treatment of maternal malaria, the safety of the artemisinin derivatives in pregnancy including in the first trimester and the need to adapt the dosing of antimalarial drugs during gestation |
| Slovakia Obstetric SurveillanceSystem (SOSS) | Slovak Obstetric Survey System (SOSS) is the working group for active surveillance of severe acute maternal morbidity and maternal mortalityin Slovak Republic (SR). It works closely with the UKOSS |
| Strengthening Epidemiological Surveillance in Benin and Burkina Fasofor an Effective Response to COVID-19(STREESCO) study | As part of the health systems set up by Benin and Burkina Faso's health authorities, this project develops with intended users, epidemic surveillance and response system that will be effective, sensitive, coordinated, and adapted to a low-resource context. The epidemic surveillance system will produce and process the ongoing information needed to execute early alerts and to control the health system's response |

| | |
|---|---|
| Sub-Saharan Congenital Anomaly Network (sScan) | support for congenital anomaly surveillance and build capacity in Sub-Saharan Africa. The network is being established by a team of investigators with experience in CA surveillance, diagnosis and care of children with CA in Africa, from different countries including; Nigeria, South Africa, Uganda and the United Kingdom |
| Swedish Medical Birth Register | The Pregnancy Register (www.graviditetsregistret.se) collects data on pregnancy and childbirth, starting at the first visit to antenatal care and ending at the follow-up visit to the antenatal care, which usually occurs at around 8–16 weeks postpartum. The majority of data is collected directly from the electronic medical records. The Register includes demographic, reproductive and maternal health data, as well information on prenatal diagnostics, and pregnancy outcome for the mother and the newborn |
| Swedish Neonatal Quality Register | SNQ provides data and information to decisionmakers, professionals, families and the public, all in order to stimulate quality improvement, research and development. Maternal, pregnancy and delivery data are automatically extracted from medical records and transferred via the Swedish Pregnancy Register |
| Swiss Neonatal Network &Follow-up group | The chief aim of the Swiss Neonatal Network & Follow-Up Group (SwissNeoNet) is to maintain and / or improve the quality and safety of medical care for high-risk newborn infants and their families in Switzerland through a coordinated program of research, education and collaborative audit. In support of its aim, SwissNeoNet hosts the official medical quality register for the Swiss level III and level IIB units. Participation for these units is mandatory according to the intercantonal declaration for Highly Specialized Medicine (HSM) of September 22, 2011 and the Society's Standards for Levels of Neonatal Care in Switzerland |
| Task Force on Research Specific to Pregnant Women and LactatingWomen (PRGLAC) | PRGLAC adviseS the Secretary of Health and Human Services (HHS) regarding gaps in knowledge and research on safe and effective therapies for pregnant women and lactating women |
| The Irish Centre for Maternal and Child Health Research (INFANT) | This is an initiative based at the University of Cork. INFANT utilizes local and global data to address the international need for research and innovation to improve health outcomes for mothers and babies |
| The PRECISE Network | The PRECISE Network is a collaboration between 13 research institutions and is hosted at King's College London in the UK, investigating three important complications of pregnancy, hypertension, fetal growth restriction, and stillbirth. |
| Ubomi Buhle | Ubomi Buhle is a national project aimed at improving our understanding of what exposures during pregnancy, such as medicines, substances and diseases, can result in poor birth outcomes e.g. birth defects, low birth weight, stillbirth, premature birth and neonatal death |
| UK Neonatal Collaborative | In 2012, the UK Neonatal Collaborative (UKNC), consisting of all NHS neonatal units, formed. These sites contribute data to the NNRD. This database, now includes details of 100,000 infants admitted to neonatal care each year |
| UK Obstetric SurveillanceSystem (UKOSS) | UK-wide surveillance network of pregnant women aimed to enable cohort, case-control, and epidemiological studies focused onrare disorders |
| US v-safe pregnancy registry | This is a registry of women who self-identify as pregnant who received COVID-19 vaccination, investigates pregnancy outcomes, pregnancy complications, and problems with the newborn |
| Vaccine Safety Datalink | The Vaccine Safety Datalink (VSD) is a collaborative project between CDC's Immunization Safety Office and nine health care organizations. The VSD started in 1990 and continues today in order to monitor safety of vaccines and conduct studies about rare and serious adverse eventsfollowing immunization. The VSD uses electronic health data from each participating site. They have a particular focus on maternal |

| | |
|---|---|
| | populations although their main focus is the general population |
| Vaccines and Medications in Pregnancy Surveillance System | The Vaccines and Medications in Pregnancy Surveillance System (VAMPSS) is a nationwide post-marketing surveillance system established to comprehensively monitor the use and safety of vaccines and medications during pregnancy |
| WHO multicountry study(WHO MCS) network | The Network for Improving Quality of Care for Maternal, Newborn and Child Health (Quality of Care Network) is a broad partnership of committed governments, implementation partners and funding agencies working to ensure that every pregnant woman, newborn and the child receives good quality care with equity and dignity. The goals of the Network are to halve maternal and newborn deaths and stillbirthsin health facilities by 2022 and to improve patients' experience of care in participating in health facilities in Network countries |
| Zero Infections in Pregnancy Honduras | Originally called the Zika in Pregancy in Honduras, this is a prospective pregnancy cohort study in Teguchigalpa, Honduras, collecting sociodemographic, maternal, and perinatal health information |
| Global vaccine data network | The GVDN addresses limitations in vaccine safety. They aim to work with vaccine safety and effectiveness experts, global health agencies, and other global non-profit health alliances to help assure the safety and risk benefit of vaccines through vaccine monitoring by evaluating vaccine safety concerns through analysis and evaluation of large clinical databases, evaluating vaccine effectiveness to facilitate risk/benefit analyses, leading a coordinated response to concerns regarding vaccines, such as vaccine hesitancy, and seeking out and securing funding for collaboration on vaccine safety monitoring projects |

**Supplementary material 4.**

**Figure 1. Results of the Quid analysis**

a) Clustering of research efforts

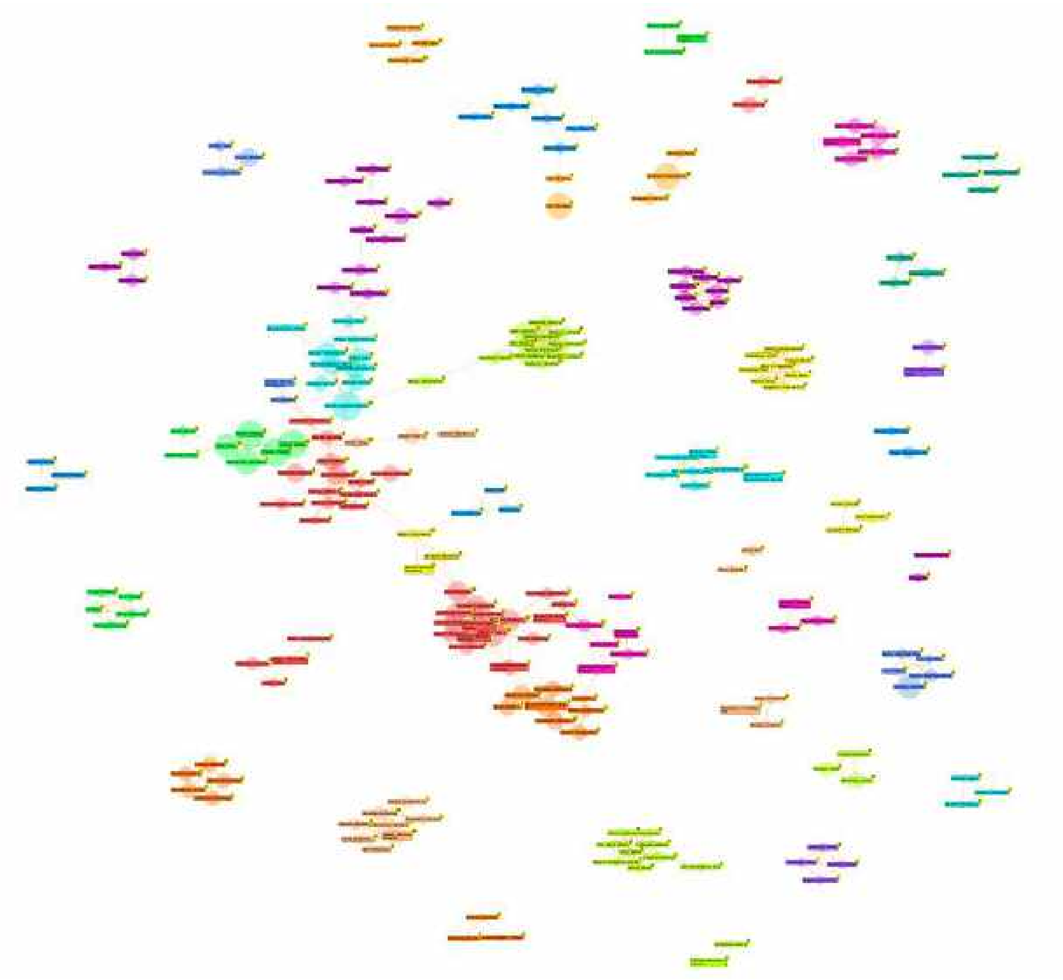

(b) focus on the most connected network

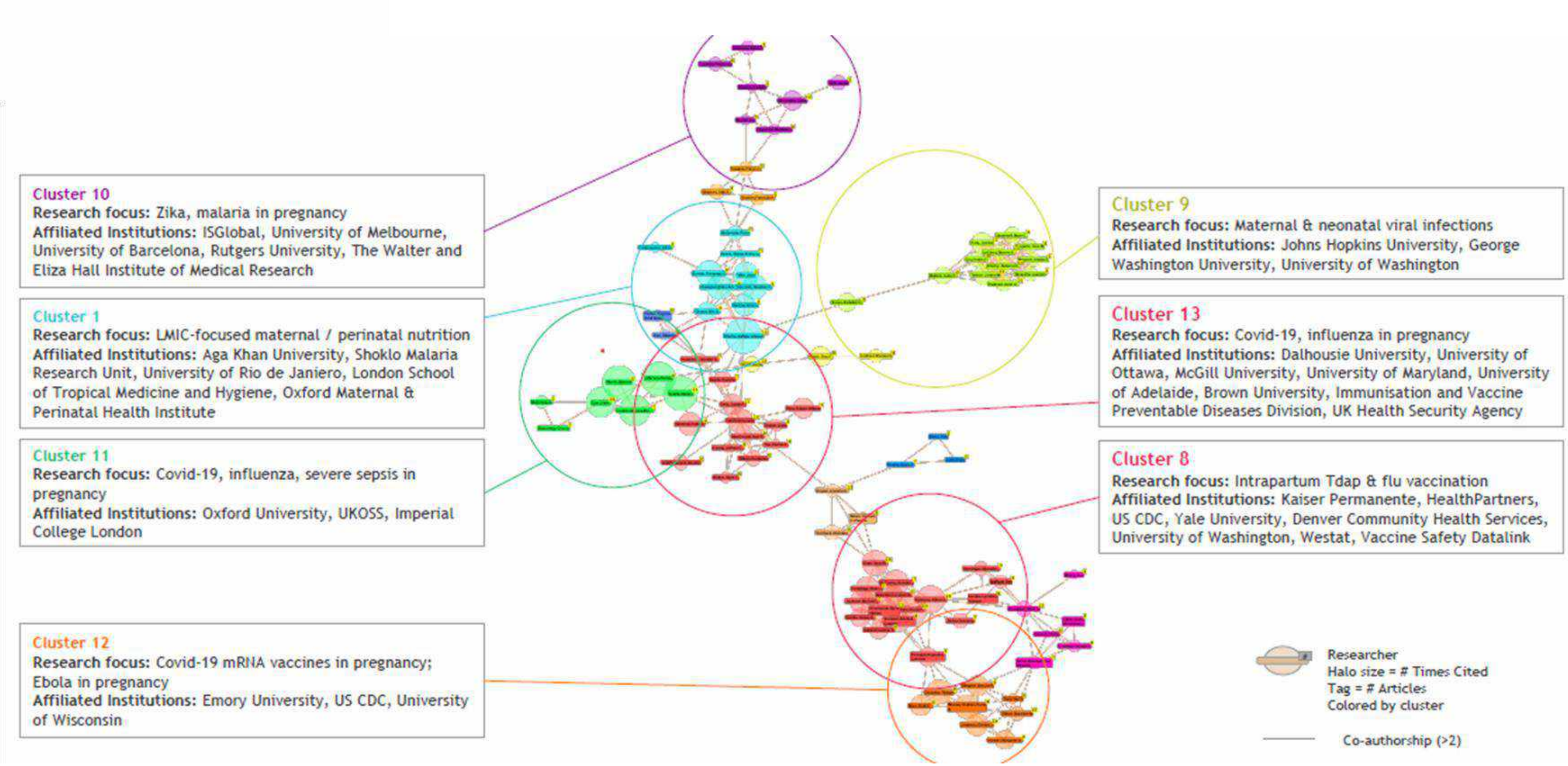

c) Maternal and perinatal health topics identified by Quid analysis

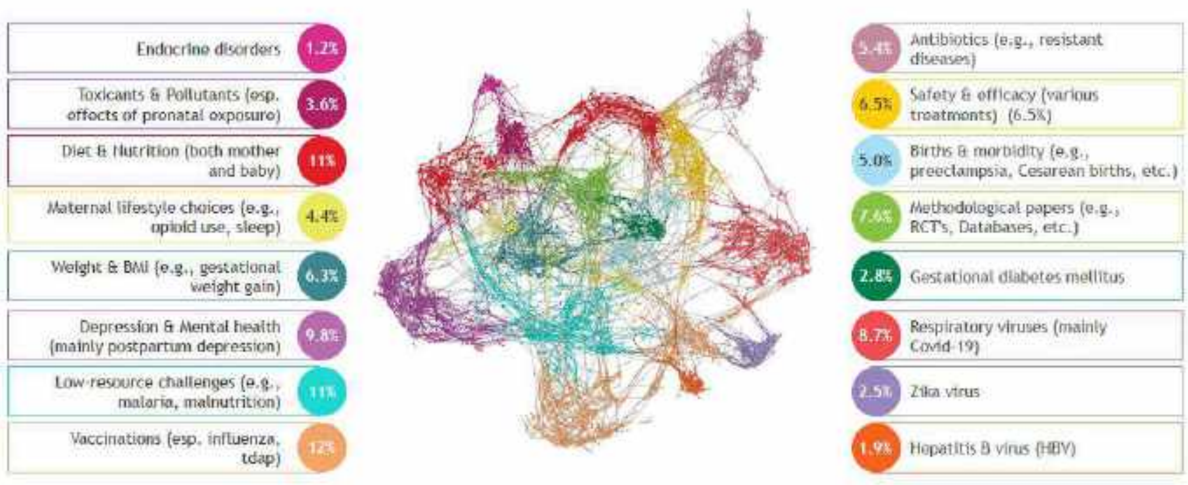
